# A comparison of various skin graft expansion models: Beyond coverage and toward improved healing

**DOI:** 10.1101/2025.10.09.25337659

**Authors:** Mohammed Asad Khan, Saiparsad Poyarekar, Vinita Puri

**Author notes:** **Corresponding author:** Mohammed Asad Khan, Indian Institute of Technology Bombay, Powai, Mumbai, India 400076.

## Abstract

The skin graft expansion techniques have demonstrated their efficacy in treating large burns in cases where the supply of donor skin is insufficient. Currently, the pursuit of skin expansion extends beyond merely covering a large area; it also aims to achieve a large perimeter gain, a smaller interpatch distance, and a shorter healing period. However, clinicians and surgeons appear to overlook the significance of inter-patch distance and perimeter gain on expansion ratio and healing period. Moreover, the interrelationship between these critical parameters is unknown to this day. This study revisits the fundamental principles of various graft expansion techniques (meshing, micrografting, and punching) to describe the interrelationship between these parameters. The analysis indicates that increasing the expansion ratio always delays the wound closure, regardless of the graft expansion technique. The recommendation provided for clinicians/surgeons to select the best surgical parameters or commercially available skin graft expansion device for improved wound healing. Furthermore, a criterion was presented to surgeons to objectively compare the two different skin graft expansion methods without bias. Furthermore, a comprehensive understanding of the interrelation between inter-patch distance, perimeter gain, wound healing period, and expansion ratio will ultimately lead to the development of novel therapeutic methods to promote large wound coverage and rapid wound healing.

## 1. Introduction

The fascinating journey of skin transplantation dates back to around 1500 BC with modest origins in ancient Egypt [1,2]. In today’s advanced era, remarkable technological progress has led to the widespread adoption of skin grafts for treating post-burn hyperpigmentation, diabetic ulcers, chronic wounds, vitiligo, and extensive burns [3–6]. Skin transplantation entails relocating skin from a specific body area (donor site) to another location (recipient site). This skin transplantation method is commonly referred to as skin grafting. Skin grafts do not possess an inherent blood supply and depend on the revascularization process from the host site for survival. The process of revascularization and reattachment of the skin graft to the recipient site is known as “graft take”. Wound healing is further driven by the proliferation and migration of the keratinocytes [7,8]. However, skin grafting has some practical drawbacks, such as creating a secondary wound equivalent to the graft size at the donor site. Even if the graft is harvested as a split-thickness skin graft (STSG), it does not entirely eradicate scarring. In addition, harvesting the skin equivalent to the wound area is impossible in the case of a larger wound area, where a healthy donor site is limited. Skin graft expansion techniques solve these inherent drawbacks of skin grafting. Skin graft expansion techniques involve obtaining grafts smaller than the wound area from a limited donor site and expanding these grafts to cover the complete wound area. Skin graft expansion techniques offer several other advantages, such as fluid drainage, facilitating optimum contact between graft and recipient bed, faster healing, reduced donor site scarring and morbidity, and the potential to avoid anesthesia and hospitalization in some instances [9,10].

In 1869, J. L. Reverdin [11] first introduced the pinch grafting method of skin graft expansion by harvesting small round grafts called “Reverdin grafts.” The pinch graft is usually harvested by pinching up the donor skin with a needle and cutting the tip of the pinch-up skin cone using a blade/scissors. The grafts obtained by pinch grafting are usually less than 5 mm in diameter, circular, deep, and contain all the skin layers. These pinch grafts are then spread over the wound bed [12]. P. Gabarro [13] first introduced the method of graft expansion by cutting tiny square pieces of graft in 1943. In this method, the graft is placed on a sticky paper and then skin with the paper cut into strips. The strips are placed on another piece of paper parallel to each other and then cut perpendicular to the strips. Finally, the cut square-shaped grafts are evenly spread over the wound bed. In 1958, Cicero Parker Meek first automated the “microdermagrafting” method using a mechanical device called the Meek-Wall microdermatome [14]. However, the micrografting technique was discontinued due to the introduction of “mesh grafting” or “meshing” by Tanner et al. [15] in 1964. The fundamental principle behind meshing involves converting a sheet graft into a lattice-like pattern and stretching to cover the wound area surgically. The primary advantage of meshing is facilitating the free fluid drainage between the graft and the recipient bed. As a result, this prevents the graft from floating over blood from the vascular bed and provides optimum contact between the graft and the underlying tissue. It also enables the immediate application of the graft to an actively bleeding surface, eliminating potential complications. Meshing a graft imparts a three-dimensional flexibility, allowing it to adapt to irregular and concave surfaces. The manual meshing of the skin graft is commonly performed if a mechanical mesher is unavailable. However, manual meshing is time-consuming and impractical for large grafts. The transfer process can cause the slits to turn into holes due to internal friction, making it nearly impossible to restore the original shape of the graft. The meshed graft becomes increasingly delicate and harder to handle at larger expansion ratios. Draping a mesh with the dermal side down onto the wound bed without damaging the mesh structure becomes progressively more difficult at larger expansion ratios. These issues associated with mesh grafting rejuvenated the modified micrografting techniques in 1993 [16]. The skin grafts produced by the micrografting are later reported by different names in literature, such as microdermagraft [14], micro skin grafts [17], skin graft particles [18], graft islets [19], pixel graft [20], seed graft [21], postage graft [16], Chip skin [22], and micrografts [23].

Over the years, numerous skin expansion devices have been developed and widely used for large areas of burns and other wounds. In addition to large wound coverage, the skin expansion graft should provide a large perimeter for epithelialization, smaller exposed (uncovered) regions, and rapid healing. Moreover, large graft expansions result in open areas of wounds, which can potentially lead to re-epithelialization delays, increased infection risks, and even graft rejection [24]. Although the significance of exposed wound area and perimeter in wound coverage and healing is crucial, it is often overlooked. To this day, it is still unknown how the spacing between the graft patches or ribbons (inter-patch distance) may affect the graft expansion. How does graft expansion affect the graft perimeter available for epithelialization? This study provides a clear relationship between surgical/design/device/operation parameters and healing parameters for various skin grafting techniques and highlights the relative importance of various parameters involved in wound coverage and healing. The surgical parameters are defined as parameters that are under the control of surgeons or device manufacturers. The surgical parameters include micrograft size, punch size, mesher blade/incision length, the vertical gap & horizontal distance between mesher blades/incisions, etc., while healing parameters describe the efficacy of the surgical process, i.e., expansion ratio, healing period, inter-patch distance, and perimeter gain. A clinician can estimate the minimum donor graft size required and the shortest wound closure time from the presented mathematical model, improving the patient’s quality of life. Furthermore, the proposed algebraic model will allow engineers to optimize current skin expansion devices and develop new, exciting techniques that could lead to rapid healing.

## 2. Materials and Methods

### 2.1 Definitions

Four healing parameters are used to describe the efficacy of a surgical procedure during skin graft expansion and healing.

#### 2.1.1 Expansion ratio (e)

The theoretical expansion ratio or stretch ratio is defined as the ratio of wound surface area to the actual donor graft area. It represents a gain in graft area due to the graft expansion process.

#### 2.1.2 Perimeter gain (PG)

The perimeter gain is the ratio of the expanded graft perimeter to the harvested graft perimeter. It represents a gain in the availability of epithelial edges due to the graft expansion process. Thus, perimeter gain represents the ability to graft an expansion device to generate new epithelial edges.

#### 2.1.3 Inter-patch distance (IPD)

The inter-patch distance signifies the distance traveled by epithelial edges during the wound healing process. The epithelial edges move inward or outward during the wound closure process. The micrografting or mini punching sites are examples where epithelial edges move outward (away from each other) to cover the targeted area. For inward-moving epithelial edges, the inter-patch distance is the perpendicular distance between the two farthest epithelial edges of the expanded graft. Meanwhile, the epithelial edges travel inward (toward each other) into the mesh skin graft site. Thus, the inter-patch distance is defined as the perpendicular distance between the two nearest epithelial edges of the expanded graft for outward-moving epithelial edges.

#### 2.1.4 Wound closure time (τ)

As the name suggests, it is the time required to cover the wound surface completely. The wound closure time can be defined as;

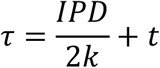

Where *k* is the rate of epithelialization and *t* is the time required for the “graft take”, which includes the revascularization and reattachment of the skin graft to the recipient site. The skin graft adheres to the recipient bed and forms a new blood supply within the time period *t*. In this article, the time required for wound closure is alternatively referred to as the wound healing period.

### 2.2 Skin graft meshing

Every mesh dermatome (mesher) comprises multiple cutting blades arranged in an alternating, staggered fashion across different rows. The mesh graft consists of alternating “full incision” and “mixed incision” rows. The “full incision” rows have each slits of length *L* while the “mixed incision” row has two *L*/2 length incisions at the end of the row. The graft expansion is achieved by pulling the skin graft perpendicularly to the length of the cuts. When tension is applied uniformly across the graft, this results in the conversion of a linear slit pattern into symmetric diamond shaped open areas. The donor skin graft has a rectangular area of sides of *X_d_* and *Y_d_*. Alternate slits of length *L* are cut on the skin graft such that each row of incision is separated from each other by a distance *D* as shown in Fig. 1 (a). The rows of incision have a vertical separation of *G*and overlap *O* such that;

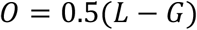

**Figure 1.**
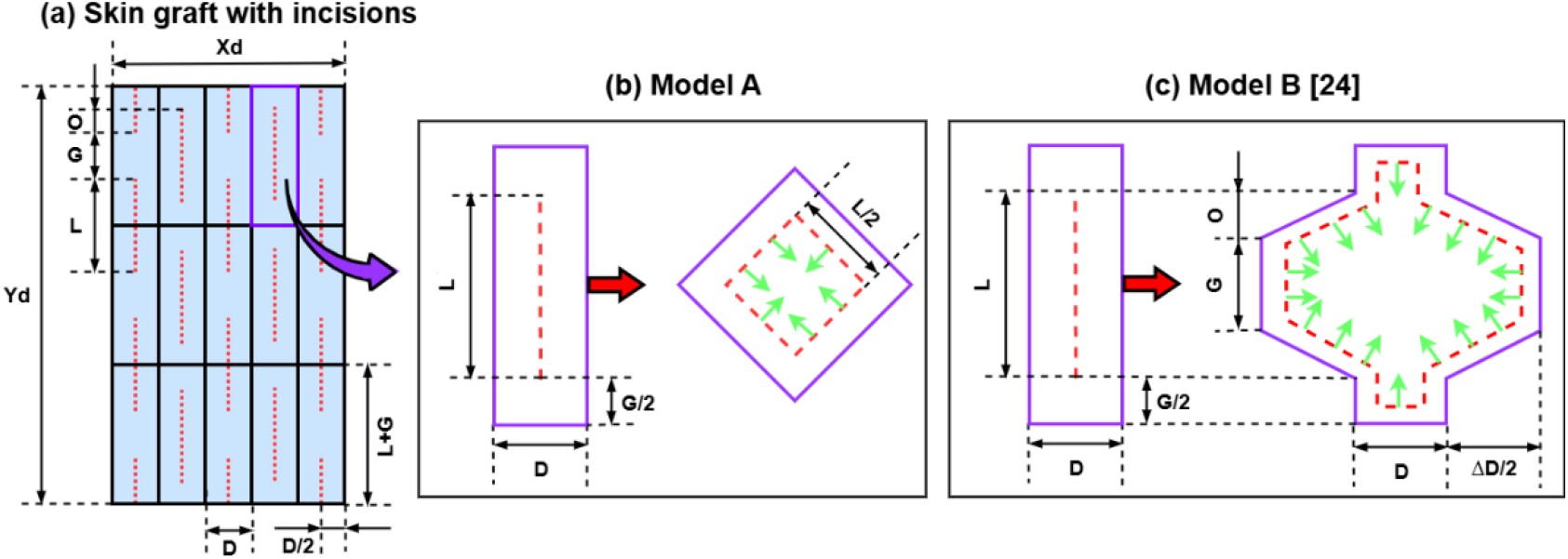
Skin graft meshing (a) Skin graft with alternating rows of interrupted incisions (red dotted lines) and effective incision area (black boxes) (b) Model A [25]: Only rotation of ribbons are considered (c) Model B [26]: Both rotation of ribbons and stretching of skin are considered (Green arrows indicates direction of keratinocytes migration)

For a mesher, *L* is the blade length, *G* is the vertical gap between blades, and *D* is the horizontal distance between blades. In some modern meshers, *L, G*, and *D* are not only dependent on the mesher blade geometry but also on the dermacarrier geometry that carries the graft into the mesher. For the sake of simplicity, *L, G*, and *D* are described as skin mesher design parameters in the present analysis. Previously, Vandeput et al. [25] presented the following relation for expanded mesh graft.

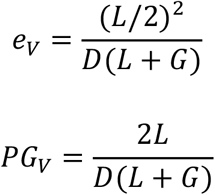

This relation was presented assuming that only the rotation of ribbons results in the expansion of the mesh graft. Also, this relation is only valid for a “fully expanded” meshed graft [27]. Vandeput et al. [25] defined a graft as a “fully expanded” meshed graft when the diamond-shaped interstices become “square-shaped interstices” as shown in Fig. 1 (b). Recently, Yu et al. [26] presented a model that captures the rotation of ribbons and the mechanical stretching of skin.

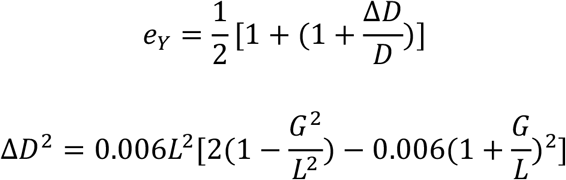

This relation is valid for under-expanded mesh with diamondshaped interstices and over-expanded mesh with dodecagonshaped interstices as shown in Fig. 1 (c). The relation is true for *L* > 0, *G*> 0, *D* > 0and *L* > *G*. The total number of “effective incision areas” or independent diamond***‐***shape areas can be calculated as follows;

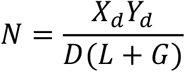

The expanded mesh graft is secured with stitches or surgical staples around the external perimeter. Therefore, only the perimeter of diamond-shaped mesh interstices practically contributes to the epithelialization process. The epithelial margins of mesh interstices release keratinocytes to spread out over the raw surface. The parameter gain indicates the available perimeter of the expanded meshed graft through which epithelial outgrowth proceeds. The perimeter gain in the expanded meshed graft is calculated relative to the original perimeter of the unmashed graft.

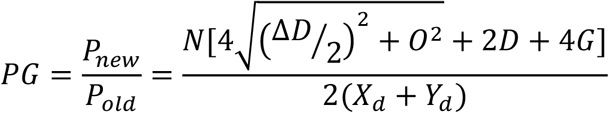

The meshed skin graft is re-epithelialized from the edges of the diamond-shaped mesh interstice towards the center. The uncovered surface area of the wound bed can be measured using the distance between the patches that cover the wound bed. The inter-patch distance is the perpendicular distance between the two nearest epithelial edges. Thus, the wound closure time of a meshed skin graft can be calculated from the inter-patch distance. The ribbons of meshed skin graft act as a patch that covers the wound bed. Therefore, the distance between the two nearest ribbons is calculated as follows;

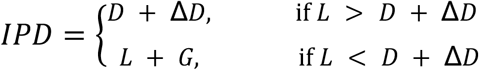

Since the whole perimeter of the diamond-shaped mesh interstice contributes to the epithelialization process, the time required for wound closure (*τ*) can be given as follows;

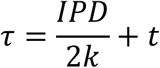

### 2.3 Micrografting

The typical micrografting process is illustrated in Fig. 2. If the donor skin graft has a rectangular area of sides of *X_d_* and *Y_d_*. The graft is then minced in perpendicular directions using a mechanical device or scissors to yield small fragments known as “micrografts” such that a single square-shaped micrograft has a side *X*_m_. Thus, the total number of micrografts obtained from a given donor area and skin graft size can be calculated as follows;

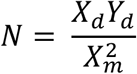

**Figure 2.**
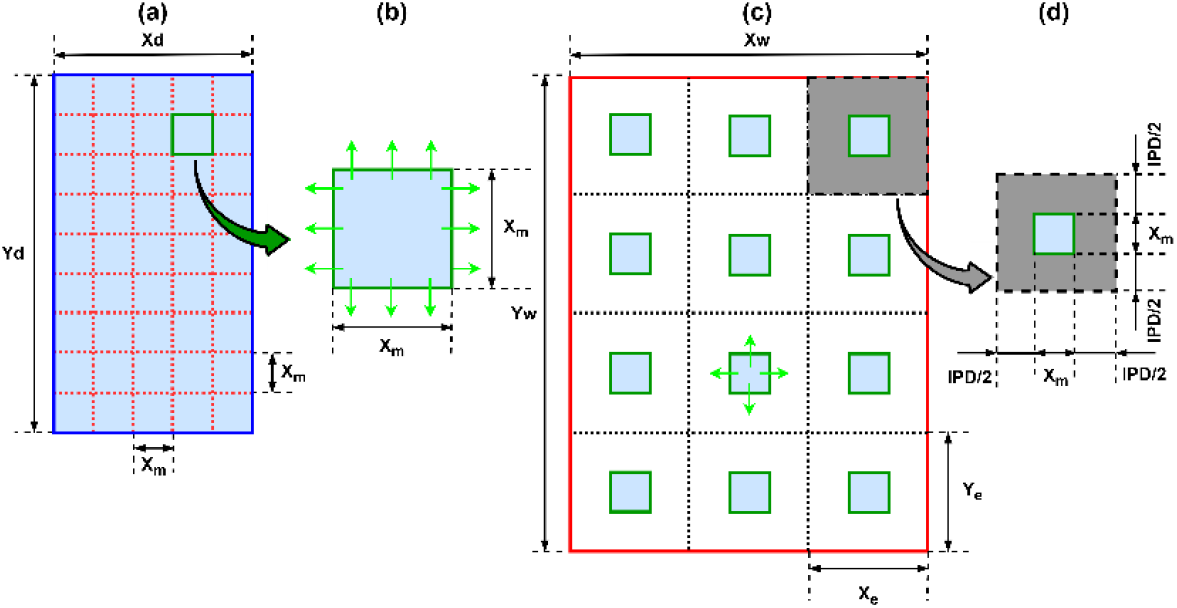
Micrografting (a) Donor skin graft (b) Single micrograft with epithelial margins (in blue) and direction of keratinocytes migration (green arrows) (c) Micrograft applied on the wound area (red lines)

The *N* number of micrografts were placed uniformly over the wound bed of *X_w_Y_w_* size, such that micrografts are parallel to each other and evenly distributed under ideal conditions. The resulting theoretical expansion ratio can be calculated as follows;

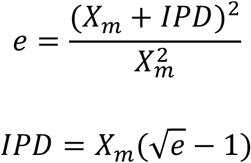

Micrografts initially survive by diffusion of wound fluid rather than neovascularisation. The proliferation and migration of keratinocytes mainly drive the healing process. The keratinocytes migrate from the graft edges to re-epithelialize the wound. Therefore, the micrografts provide more active edges for regeneration compared to a single large graft. The perimeter gain due to micrografting compared to the original skin graft is calculated as follows;

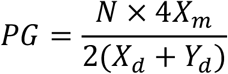

The micrograft is re-epithelialized from the edges outward direction. The micrografts act as a patch that covers the wound bed. Therefore, the time required for wound closure (*τ*) can be given as follows;

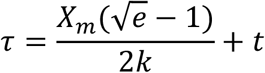

### 2.4 Mini punch grafting

If the circular punch grafts of diameter *D_m_* be taken from a donor area of *X_d_Y_d_* with the help of a punch and scissor or suction blister device.

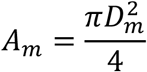

Since the donor graft is harvested in the form of mini punches instead of taking the whole donor area as shown in Fig. 3. Thus, the effective donor graft area for the *N* number of punches is calculated as follows;

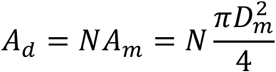

**Figure 3.**
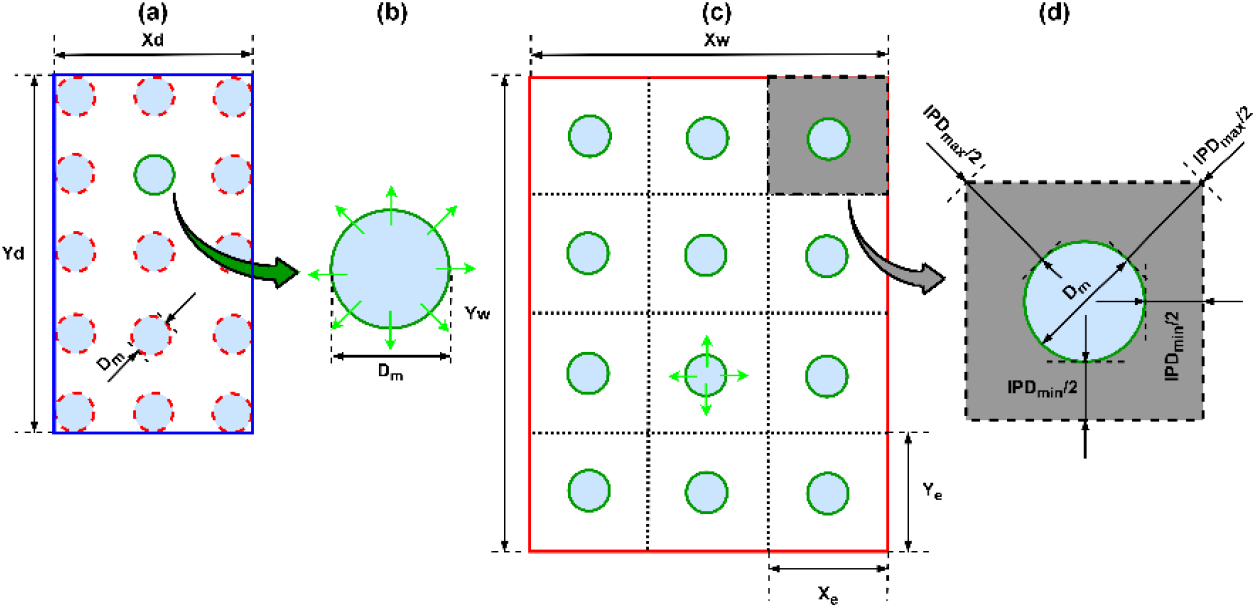
Mini punch grafting (a) Donor skin graft (b) Single mini punch graft with epithelial margins (blue) and direction of keratinocytes release (green arrows) (c) Mini punch graft applied on the wound area (red lines)

The *N* number of punch grafts gently spread over the abraded vitiliginous area of size *X_w_Y_w_*. The resulting theoretical expansion ratio can be calculated as follows;

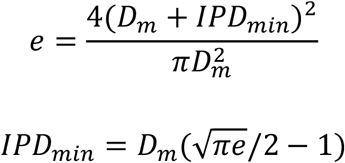

These expressions are valid only for *e* > 4/*π*. In 1972, Norman Orentriech [28] first extended the pinch graft method for treating long-term leucoderma after a chemical burn. He deployed several circular normal skin autografts of a few millimeters in diameter and observed the “pigment spread phenomenon”. Epithelialization starts first, and repigmentation begins a few weeks after epithelialization is complete. The repigmentation occurs through the proliferation of melanocytes and the spreading of pigment from grafts. Active and functional melanocytes migrate centrifugally from the edge of the punched graft to recolonize the area [29]. The individual punch should replenish the wound area. The two nearest circular mini punches have a maximum inter-patch distance as follows.

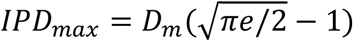

The perimeter gain and time required for wound closure (*τ*) for mini punch grafting can be written based on maximum inter-patch distance as follows.

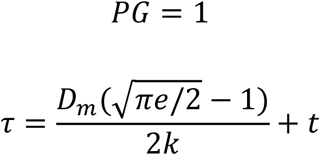

## 3. Resutls and Discussions

### 3.1 Influence of mesher design parameters on healing parameters

Figure 4 (a) compares the real and claimed expansion of the mesh graft. It is well-known that the expansion ratio obtained during clinical practice is usually different from that claimed by the manufacturers [30–33]. The claimed expansion ratios (*e_c_*) are generally much higher than the true or real expansion ratios (*e_R_*). The theoretical expansions calculated from Model A and Model B are also plotted in Fig.4 (a). It is important to note that Model A significantly overestimates the expansion ratio. Meanwhile, the Model B accurately predicts the expansion ratio, which can be seen by an overlapping curve of the real expansion (black line) and the Model B expansion (Green line). The accurate theoretical prediction of mesh graft expansion ratios from the Model B is expected, as it combines the effects of ribbon rotation and skin stretching. Figure 4(b) shows a negative correlation between the percentage of expected expansion achieved in practice for different claimed expansion ratios. The analysis shows that the projected expansion of donor grafts can be achieved when targeted expansion ratios are small. As the expansion ratio becomes larger, the percentage of expected expansion decreases. A similar observation was reported by Richard et al. [31]. The comparison of inter-patch distance and percentage of expansion achieved for various skin meshing devices is tabulated in Appendix A. The skin mesher design parameters (*L, G*, and *D*) and claimed expansion ratio (*e_c_*) provided by the manufacturer were obtained from Vandeput et al. [37,38]. The real expansion ratio (*e_R_*) is approximated using Model B. If the epithelialization rate (*k*) is assumed to be 1 mm per day, then the time required for wound closure (*τ*) is estimated as *τ* = 0.5*IPD* by ignoring the time required for revascularization (*t* = 0). It shows that all different brands of meshers have failed to achieve the expected expansion ratio. No mesher brand appeared as a winner among different meshing systems in terms of the percentage of expected expansion achieved in practice. All meshing systems perform in similar ways and display a negative correlation between the percentage of expansion and claimed expansion ratios. At a lower expansion ratio (*e_c_* < 2), all meshing systems provide larger than 60% of the expected expansion. The percentage of expected expansion decreases for all meshers as the expansion ratio increases. The mesher gives lower than 50% of the expected expansion for expansion ratios (*e_c_*) larger than 4. For all commercially available meshing systems, the blade length (*L*) is much higher than the horizontal distance between blades (*D*). Thus, the *IPD* of meshers are calculated as *D* + *δD*.

**Figure 4.**
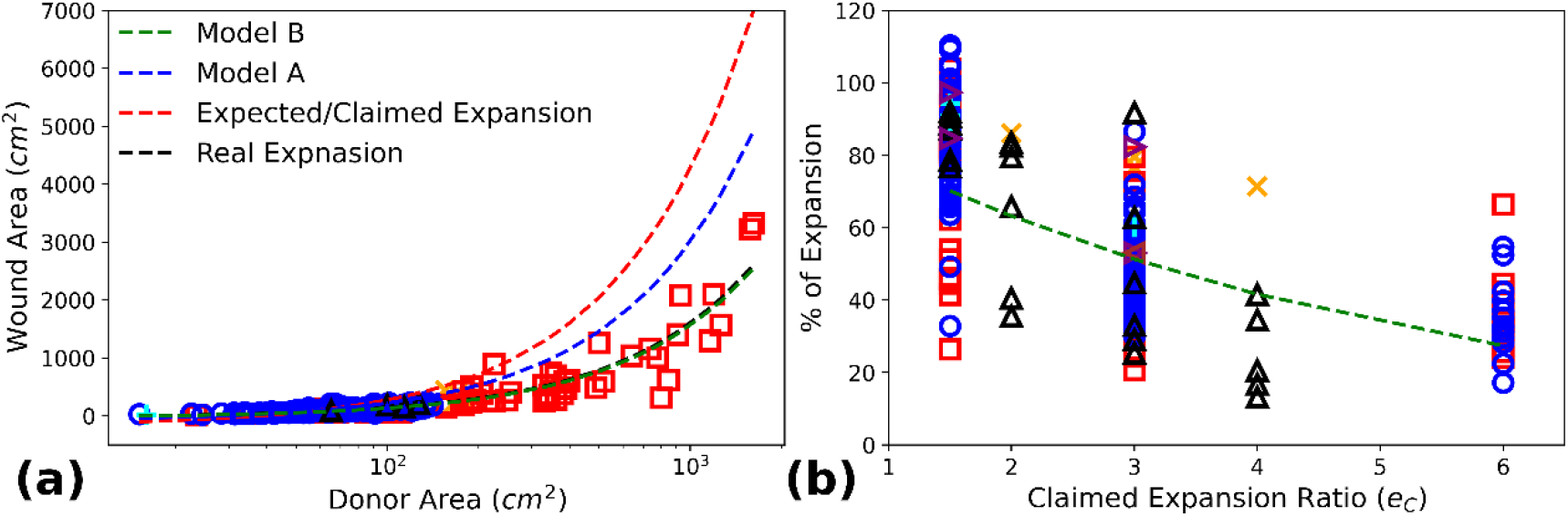
(a) Real expansion 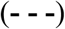and claimed 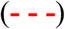expansion from the literature Kamolz et al.[30] 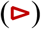, Richard et al.[31] (**△**), Lyons et al.[32] 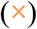, Kan et al.[33] 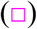, Lumenta et al.[34] 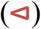, Henderson et al.[35] 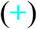, Peeters et al.[36] 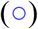compared with theoretical expansion calculated from Model A [25] 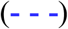and Model B [26] 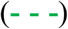 (b) Percentage of real expansion achieved for different claimed expansion ratios

The present analysis answers the basic question of how design parameters influence wound healing parameters using the new Model B. The Model B is selected since it accurately predicts the real expansion ratios and is valid for all mesh interstice shapes (diamond, square, dodecagon). The effect of mesher design parameters on expansion ratio, inter-patch distance, and perimeter gain for different expansion ratios was explained using Fig. 5. The values are plotted for a donor area equivalent to an A4-size paper (∼20mm × 30cm). The mesher design parameters are selected as *L* = 110mm, *G*= 3mm, and *D* = 2mm unless they are the variables plotted along the x-axis. All mesher design parameters (*L, G*, and *D*) play a key role in defining the expansion ratio, inter-patch distance, and perimeter gain. The main objective of skin graft expansion is to gain the highest possible expansion ratio. The meshed grafts can achieve a higher expansion ratio using the large values of *L* and lower values of *D* and *G*. However, among the mesher design parameters, the *D* has the highest influence on the expansion ratio compared to *L* and *D*. This means the smallest changes in *D* result in huge variations in the expansion ratio. The *L* and *G* have moderate and the lowest effect on the expansion ratio, respectively. For skin graft expansion, the smallest inter-patch distance is recommended since *IPD* directly affects the time required for wound closure. The lower *IPD* for mesh graft can be maintained with a large value of *G*and lower values of *L* and *D*. The influence of *D* and *L* is strongest and moderate on *IPD*, respectively. Meanwhile, the *G*has marginal effect on *IPD*. The analysis shows that the effect of *L* and *G*is weak and opposite on the expansion ratio and *IPD*. It means higher (lower) values of *L* (*G*) give a large expansion ratio but also result in a large inter-patch distance, which prolongs the wound closure period. Furthermore, the *D* emerges as the optimal parameter that has the strongest influence on the expansion ratio and inter-patch distance, such that a lower value of *D* favors the desired objective of the highest expansion and the lowest inter-patch distance. Therefore, clinicians and manufacturers should select values of *D* as small as possible to ensure the large graft expansion with rapid wound closure. On the other hand, large perimeter gain can be obtained for the lower values of *L, G*, and *D*. The perimeter gain is mainly affected by *D* and is weakly affected by *G*. Till now, this novel interrelation between different mesher design parameters and healing parameters has not been reported in the literature.

**Figure 5.**
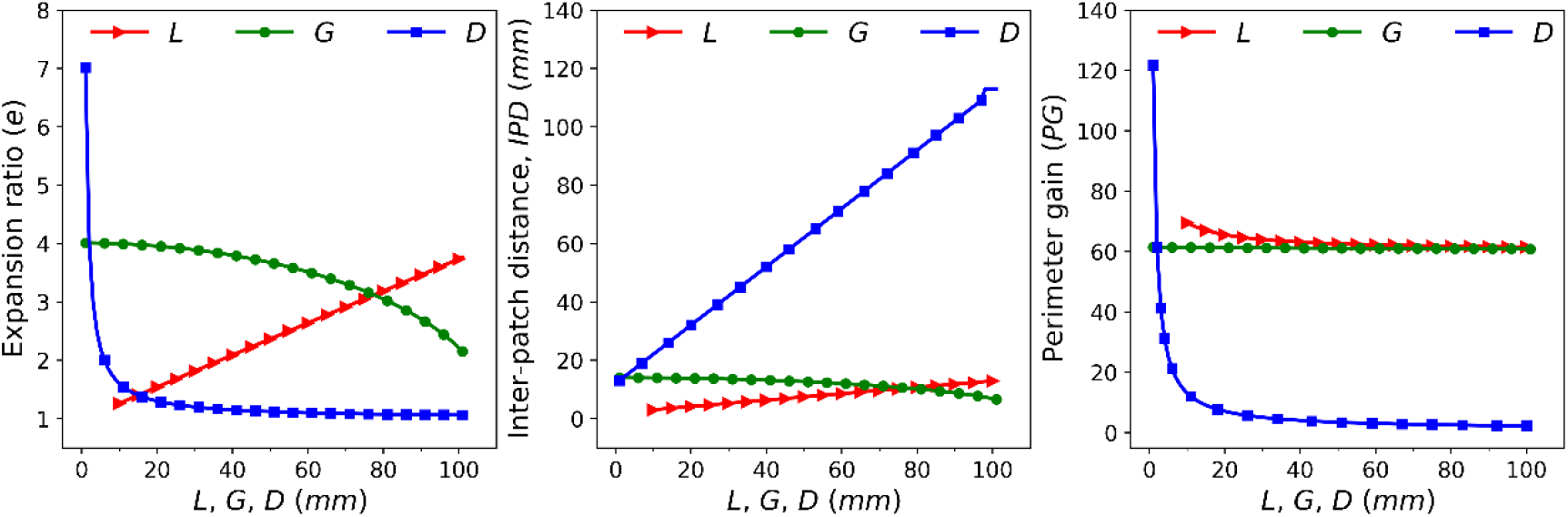
The effect of mesher design parameter (*L, G*, and *D*) on expansion ratio (e), inter-patch distance (*IPD*) and perimeter gain (*PG*)

### 3.2 Effect of micrograft size and expansion ratio

Figure 6 compares the proposed model with the previous model presented by Lin et al.[39]. Lin’s model overestimates the inter-patch distance and total healing time since Lin’s model also accounts for the diagonal distance between two nearest micrografts. The presented new model only considers a perpendicular distance between the two closest epithelial edges, which are mainly responsible for keratinocyte migration, re-epithelialization, and overall wound closure. The accuracy of the presented model can be validated by calculating the inter-patch distance for expansion ratio *e* = 1. Lin’s model gives a non-zero value of inter-patch distance while the actual value should be *IPD* = 0 since there is no expansion of skin graft at *e* = 1. Figure 6 shows the effect of micrograft size and expansion ratio on the *IPD* and perimeter gain for the A4 size wound area. The algebraic model shows that inter-patch distance and, therefore, wound healing time linearly increase with micrograft size. Similarly, the expansion ratio increases the healing time by increasing the inter-patch distance. However, the *IPD* or healing time is significantly affected by the micrograft size compared to the expansion ratio. The model also illustrates that the perimeter gain exponentially decreases with the micrograft size. Therefore, the best practice is to reduce the micrograft size as small as possible, such that the highest expansion ratio with the lowest inter-patch distance can be obtained. It is interesting to note that the perimeter gain is independent of the expansion ratio. Thus, the perimeter gain should be used to compare the effectiveness of micrografting devices instead of the expansion ratio. A micrografting techniques that provide large perimeter gain would necessarily result in rapid wound closure compared to a device that gives smaller perimeter gain, if the expansion ratio of both devices is equal.

**Figure 6.**
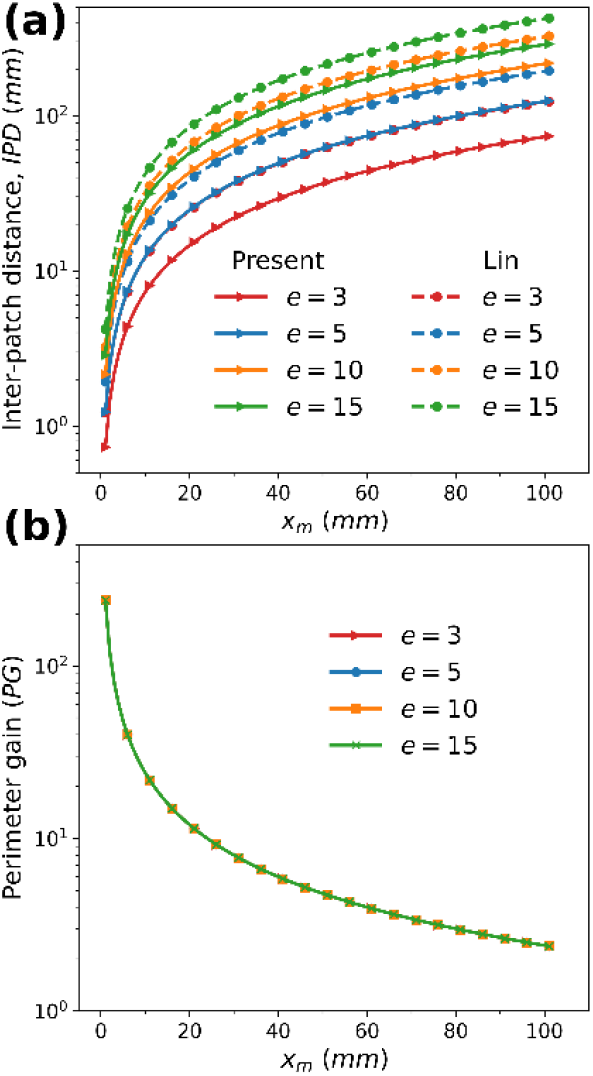
(a) Comparison of Lin et al.[39] and present model for inter-patch distance (*IPD)* (b) The relation between micrograft size (*X_m_*), expansion ratio (e), and perometer gain (*PG*)

Kreis et al.[16] reintroduced a modified Meek technique that divides pieces of split skin grafts into smaller rectangular skin graft islands. The Humeca MEEK device has 13 parallel, round blades spaced 3mm apart from each other. The blades cut the 42mm × 42mm graft into 14 strips in two perpendicular directions, resulting in a total of 196 square micrografts of 3mm size. The folded polyamide gauze with aluminum foil backing generates graft expansion ratios of 1:3, 1:4, 1:6, and 1:9 [40]. Similarly, the Xpansion® mincing system consists of 24 parallel rotating cutting disks 0.8mm apart. The resulting micrografts have a 0.8mm size[20]. Appendix B compares the inter-patch distance and perimeter gain of MEEK and Xpansion® devices for different expansion ratios for a donor area equal to A4 paper size. The Xpansion® is the clear winner in terms of larger perimeter gain and shorter healing time due to the smallest micrograft size. However, it is quite difficult to obtain uniformly distributed micrografts over a wound surface with Xpansion®. In contrast, prefolded gauze in MEEK devices allows a uniform distribution of micrografts with constant spacing. This practical limitation of Xpansion® might increase the total duration of wound healing.

### 3.3 Role of punch size and expansion ratio

Figure 7 shows the relationship among mini punch size (*D_m_*), expansion ratio (e), and inter-patch distance (*IPD*). The larger punch size and expansion ratio extend the healing period by increasing the inter-patch distance. The model also shows the linear relationship between the punch size and inter-patch distance. In addition, punch size has a dominating effect on inter-patch distance compared to expansion ratio. Therefore, it is recommended to use the smallest possible punch size to get the highest expansion with rapid wound healing. Several punches, i.e., Keye’s punch, loo trephine, disposable punch, etc., are available commercially in sizes ranging from 1mm to a few centimeters. In addition, specialized devices that work on the prolonged suction of the donor site at negative pressure are available in the global market. A suction blister device, Dermovac®, consists of suction cups and a hand pump. The suction cups can produce 5, 2, and 17 suction blisters of 4mm, 3mm, and 1.5mm, respectively [41,42]. Similarly, the Korean/Chinese suction cups with valves have suction blisters of 20 to 25 mm in size [43,44]. In literature, blisters can also be generated using syringes with 2ml, 5ml, 10ml, 20ml, and 50ml capacity and 3-way connectors [44–46]. The recently developed CelluTome™ epidermal harvesting system automatically applies heat and suction to the donor site. The CelluTome™ harvester produces 128 suction blisters of 2mm size separated from each other at 2 _mm_ and spread over an area of 2.5cm × 2.5cm or 5cm × 5cm. It also allows the epithelial graft to be quickly transferred to the recipient site with wound gauze. As a result, the CelluTome™ has been shown to reduce procedure time and minimize discomfort [47–49]. The inter-patch distances for various commercial punch grafting systems for a given expansion are tabulated in Appendix C. The in-house specialized suction blister devices used by several clinicians are not included.

**Figure 7.**
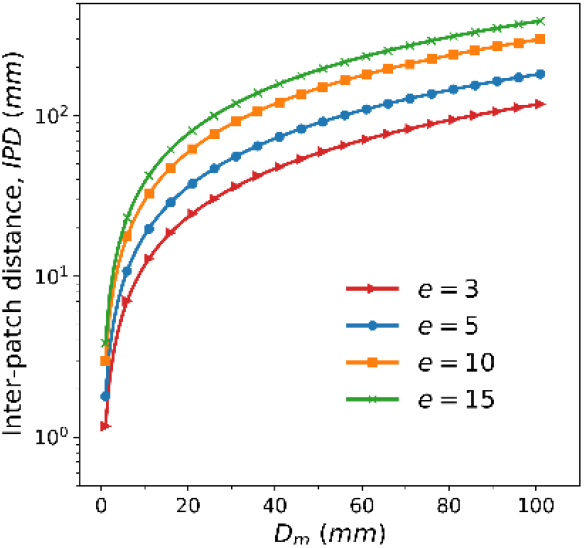
The effect of mini punch size (*D_m_*) and expansion ratio (e) on inter-patch distance (*IPD*)

### 3.4 Comparison of meshing, micrografting, and punching

The comparison of different types of skin graft expansion techniques, as well as commercially available devices, shows that there is a trade-off between the expansion ratio and *IPD* or time required for wound healing. A skin grafting tradeoff is a situation in which graft expansion to cover a large wound area can’t be achieved without compromising rapid wound closure or increasing the distance between the epithelial edges (*IPD*). In other words, the higher expansion ratio can be achieved only at the expense of the time required for wound healing. Figure 8 compares the interpatch distance and perimeter gain of different skin graft expansion techniques over a range of expansion ratios. The values are plotted for a donor graft area equal to A4 paper size. The horizontal distance between the mesher blade (*D*), micrograft size (*X_m_*), and punch diameter (*D_m_*) are the key parameters to obtain optimal expansion ratio and rapid wound healing as discussed in the above sections. Thus, these three key parameters are assumed constant and equal to 0.5 mm. The other two mesher parameters are estimated from the Model B for a given e and *D*. The inter-patch distance increases directly as the square root of the expansion ratio for micrografting and mini-punch grafting. Meanwhile, the inter-patch distance of the meshed graft increases linearly with the expansion ratio. The strongest trade-off between the graft expansion and wound healing period was observed for meshing. The micrografting shows the weakest tradeoff between expansion ratios and inter-patch distance among all three skin grafting techniques. Similarly, the perimeter gain is higher for micrografting compared to meshing. Moreover, the perimeter gain is constant at the lowest value of 1 for minipunching. Therefore, the micrografting technique emerges as the winner in terms of lower inter-patch distance and highest perimeter gain, irrespective of expansion ratio. Figure 8 compares the expansion ratio and perimeter gain required by different expansion techniques for a range of inter-patch distances or wound closure times. The graph is plotted for 0.5 mm fixed values of mesher blade (*D*), micrograft size (*X_m_*), and mini punch diameter (*D_m_*). The *L* and *G* are calculated from *D* and *IPD* using Model B. From the comparison, micrografting emerges as a technique that gives maximum graft expansion for the given wound healing period. In other words, the micrografting method requires a minimum donor graft area for a given wound size and wound healing period. The *D* for meshing should be 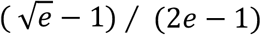times *X_m_* and 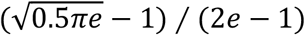 times *D_m_* to provide the same expansion ratio and wound closure time as micrografting and punching, respectively. The meshing will always have a higher wound closure time compared to meshing if 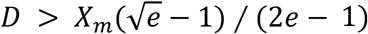 Dahmardehei et al.[50] compared the meshing and micrografting methods in a control study on 20 third-degree burn patients with 4 and 6 expansion ratios. The mesh dermatome of *D* = 1.27 mm and MEEK device of *X_m_*= 3 mm was used, which shows mean repithelialization time ± 2.5 months and 5.0 ± 2.1 months for micrografting and meshing, respectively. The *D* for the clinical study is higher than 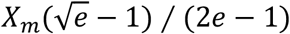 and therefore micrgrafting technique results in rapid wound closure compared to meshing be. Thus, the expansion ratio as well as the inter-patch distance should be equal to perform a comparative study of two different graft expansion techniques. Moreover, the algebraic models also show that the perimeter gain obtained from micrografting is the highest among all three techniques. In summary, the analysis shows that micrografting performs best in all parameters compared to meshing and mini punching.

**Figure 8.**
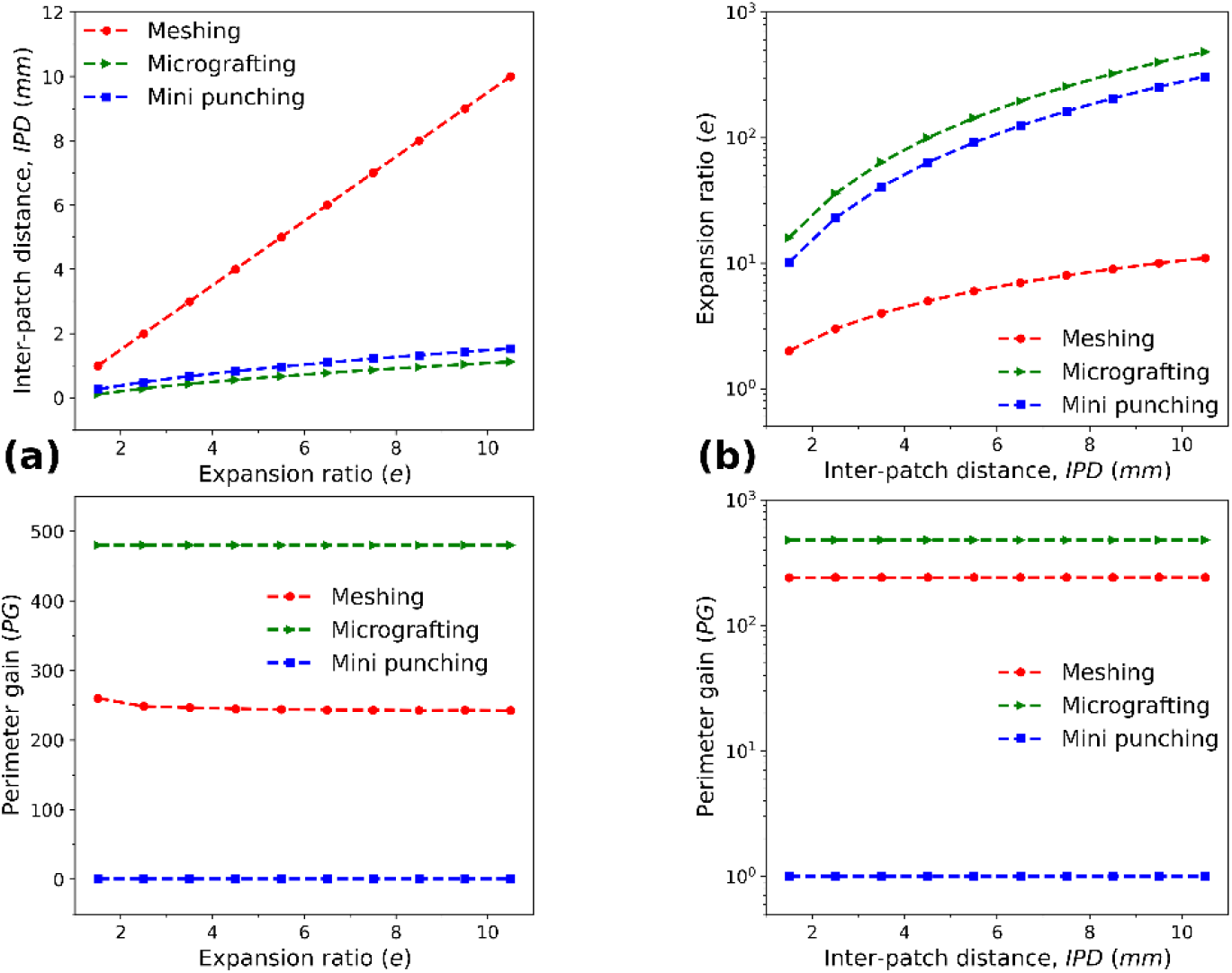
Comparison of meshing, micrografting, and punching (a) for different expansion ratios (b) for different inter-patch distances or wound healing time

### 3.5 Limitations of the presented approach

A few limitations of the presented approach need to be acknowledged. In this analysis, the proposed mathematical relationship is established for an ideal condition based on certain assumptions and simplifications, which may not fully capture the complexity of the skin grafting procedure. The recipient area is approximated as rectangular in shape, in which micrografts were correctly orientated and uniformly distributed. The role of micrografts or punch clusters that were observed in real scenarios was not accounted for in this model. Moreover, it is assumed that all edges of the donor graft contribute to the epithelialization process. Also, the epithelialization rate (*k*) was assumed to be constant for all micrografts and each boundary of the individual micrograft. In actual practice, the micrograft or punches are difficult to disperse over the wound with a fixed inter-patch distance, except in some devices. Nevertheless, the mathematical models give a better understanding of the relationship between various parameters and provide a comparative framework for various skin grafting techniques.

## Conclusion

The basic principles of skin grafting are reviewed, and more accurate algebraic models are presented for micrografting and punching techniques. The proposed models describe the relation between various parameters essential for rapid wound closure and healing. A novel interrelation between different mesher parameters and healing parameters has been reported. The model further highlights the importance of inter-patch distance in reducing the healing period and days of hospital stay. The mathematical analysis presented in this paper is summarized as follows.

1. The horizontal distance between the blades or space between two incision rows (*D*) emerges as an optimal parameter that should be as small as possible for maximum mesh graft expansion and rapid wound healing.
2. Among the different brands of meshers available in the market used in the present study, no single brand is a winner in terms of achieving the expected expansion ratio.
3. It is recommended that the first row of mesh graft be a “full incision” row and a total odd number of rows, especially during manual preparation of mesh grafts. As a result, a surgeon is required to make the lowest number of incisions without affecting the expansion ratio. Also, the boundary of the final mesh will be less damaged due to open interstices. This makes manual meshing less labour-intensive and facilitates easy transfer of the final expanded mesh graft on the recipient site.
4. The micrograft size (*X_m_*)/punch size (*D_m_*) is a key parameter that strongly influences the expansion ratio and inter-patch distance in the context of micrografting or punching.
5. The algebraic models reveal that aiming for a large expansion ratio due to the unavailability of donor sites always prolongs the healing period, with the possibility that healing may not occur at all. This tradeoff between graft expansion and wound healing time exists for all graft expansion techniques.
6. Micrografting manifests the lowest degree of trade-off between graft expansion and healing time compared to meshing and mini-punch grafting. It means the highest wound coverage per unit donor area can be yielded at the expense of wound closure time, and vice versa with the micrografting technique. Thus, microfracturing appears superior skin grafting method among all graft expansion techniques.
7. A criterion was presented to compare the two different skin graft expansion techniques. For comparison of two methods, the expansion ratio as well as the inter-patch distance of the two methods should be equal. Otherwise, the method with a lower interpatch distance always results in rapid reepithelialization with the shortest wound closure time.

The proposed algebraic relations serve as a guideline for clinicians in selecting appropriate techniques, optimum donor graft size, estimating the wound healing period, total hospitalization time, etc. Moreover, the comprehensive understanding of various skin grafting principles and the algebraic relationship presented here will also pave the way for innovative skin grafting and expansion approaches. The future work would be to develop an open-source digital tool for the calculation of the donor graft size and other healing parameters

## Data Availability

All data produced in the present study are available upon reasonable request to the authors

## Notes

### Competing Interest Statement

The authors have declared no competing interest.

### Funding Statement

This project has received a grant from the Government of India IDEX-DIO, DBT-BIRAC,
and MeitY programs. The authors also acknowledge CSR support from SINE IIT Bombay,
ICICI Securities, CITI Bank, and HDFC Bank.

