## Appendices for "A comparison of various skin graft expansion models: Beyond coverage and toward improved healing"

**Appendix A** Comparison of inter-patch distance and percentage of expansion achieved using various skin meshing apparatus (All units are in mm)

| **Sr. No.** | **Device name** | $\boldsymbol{L}$ | $\boldsymbol{G}$ | $\boldsymbol{D}$ | $\boldsymbol{e}_{\boldsymbol{C}}$ | $\boldsymbol{e}_{\boldsymbol{R}}$ | $\boldsymbol{\%}$ | $\boldsymbol{IPD}$ |
| --- | --- | --- | --- | --- | --- | --- | --- | --- |
|  | Aesculap | 8.13 | 0.41 | 1.52 | 3.00 | 1.29 | 43.04 | 2.41 |
|  |  | 18.29 | 10.16 | 1.52 | 6.00 | 1.54 | 25.73 | 3.18 |
| 3 | Ampligreffe (Collin) | 7.62 | 3.05 | 1.02 | 2.00 | 1.38 | 68.76 | 1.78 |
|  |  | 15.24 | 3.05 | 1.02 | 4.00 | 1.80 | 45.08 | 2.65 |
|  |  | 22.35 | 3.05 | 1.02 | 6.00 | 2.19 | 36.52 | 3.44 |
|  |  | 30.48 | 3.05 | 1.02 | 8.00 | 2.63 | 32.90 | 4.33 |
| 4 | Brennen, Bioplasty, Padgett | 5.51 | 2.82 | 1.02 | 2.00 | 1.25 | 62.71 | 1.53 |
|  |  | 8.38 | 2.82 | 1.02 | 3.00 | 1.43 | 47.48 | 1.88 |
|  |  | 13.97 | 0.28 | 1.02 | 4.00 | 1.75 | 43.79 | 2.54 |
|  |  | 30.99 | 0.28 | 1.02 | 6.00 | 2.67 | 44.47 | 4.41 |
|  |  | 48.01 | 0.28 | 1.02 | 8.00 | 3.59 | 44.81 | 6.27 |
| 5 | Zimmer® Mesh-dermatome II | 4.80 | 2.47 | 1.27 | 1.50 | 1.18 | 78.45 | 1.72 |
|  |  | 10.16 | 5.64 | 1.27 | 3.00 | 1.36 | 45.42 | 2.19 |
|  |  | 24.13 | 14.35 | 1.27 | 6.00 | 1.83 | 30.53 | 3.38 |
|  |  | 43.79 | 22.53 | 1.27 | 9.00 | 2.61 | 29.02 | 5.36 |
| 7 | Zimmer® Skingraft mesher | 4.57 | 0.76 | 1.27 | 1.50 | 1.19 | 79.60 | 1.76 |
|  |  | 7.87 | 0.76 | 1.27 | 2.00 | 1.34 | 66.87 | 2.13 |
|  |  | 10.16 | 0.76 | 1.27 | 3.00 | 1.44 | 47.88 | 2.38 |
|  |  | 15.24 | 0.76 | 1.27 | 4.00 | 1.66 | 41.39 | 2.93 |
| 8 | Zimmer® Mesh-dermatome I | 7.62 | 0.76 | 1.27 | 3.00 | 1.33 | 44.27 | 2.10 |

**Appendix B** Comparison of inter-patch distance and perimeter gain for various skin micrografting devices

| **Device name** | $\boldsymbol{X}_{\boldsymbol{m}}$**(mm)** | $\boldsymbol{e}$ | $\boldsymbol{IPD}$ **(mm)** | $\boldsymbol{PG}$ | $\boldsymbol{\tau}$ **(days)** |
| --- | --- | --- | --- | --- | --- |
| Humeca MEEK | 3.00 | 2.00 | 1.24 | 80 | 3.00 |
|  | 3.00 | 3.00 | 2.20 | 80 | 3.00 |
|  | 3.00 | 4.00 | 3.00 | 80 | 3.00 |
|  | 3.00 | 6.00 | 4.35 | 80 | 3.00 |
|  | 3.00 | 9.00 | 6.00 | 80 | 3.00 |
| Xpansion® | 0.80 | 2.00 | 0.33 | 300 | 0.80 |
|  | 0.80 | 3.00 | 0.59 | 300 | 0.80 |
|  | 0.80 | 4.00 | 0.80 | 300 | 0.80 |
|  | 0.80 | 6.00 | 1.16 | 300 | 0.80 |
|  | 0.80 | 9.00 | 1.60 | 300 | 0.80 |

**Appendix C** Comparison of inter-patch distance for various commercially available mini punch grafting techniques (All units are in mm)

| **Device name** | $\boldsymbol{D}_{\boldsymbol{m}}$ | **No. of punch** | $\boldsymbol{e}$ | $\boldsymbol{IPD}$ |
| --- | --- | --- | --- | --- |
| CelluTome™ epidermal harvesting system | 2.0 | 128 | 5.10 | 3.66 |
| Dermavac® | 4.0 | 5.0 | 5.10 | 7.32 |
|  | 3.0 | 2.0 | 5.10 | 5.49 |
|  | 1.5 | 17 | 5.10 | 2.74 |
| Korean/Chinese suction cups | 20-25 | 1.0 | 5.10 | 36.59 - 45.74 |
| Syringes with 3-way connectors | 10.00 | 1.0 | 5.10 | 18.30 |
|  | 14.00 | 1.0 | 5.10 | 25.62 |
|  | 17.00 | 1.0 | 5.10 | 31.10 |
|  | 22.00 | 1.0 | 5.10 | 40.25 |
|  | 32.00 | 1.0 | 5.10 | 58.55 |
